# Neurodevelopmental Vulnerability in Cognitive Aging: A Dimensional Approach

**DOI:** 10.64898/2026.01.15.26343771

**Authors:** Perrine Laury Marie Siguier, Mélanie Planton, Bérengère Pages, Marie Wolfrum, Ombeline Archambault, Anise Damour, Valentine Guidolin, Pauline Pefferkorn, Lola Danet, Laurine Virchien, Eloi Magnin, Aurélie Richard-Mornas, Mathilde Sauvée, Catherine Thomas-Antérion, Servane Mouton, Jérémie Pariente, Mélanie Jucla

## Abstract

Neurodevelopmental disorders (NDDs) influence lifespan neurocognitive trajectories and can be conceptualized as falling on a continuum. However, transdiagnostic neurodevelopmental investigations in cognitive aging are rare. This preliminary, cross-sectional study aimed at exploring lifespan manifestations of neurodevelopmental vulnerabilities (DVs) in cognitive aging, while adopting a dimensional approach to NDDs. The objectives, covered from childhood to adulthood, were: 1) to describe NDDs-related domains of vulnerability; 2) to estimate DVs frequency; 3) to evaluate the persistence of DVs symptoms from childhood to adulthood; 4) to explore the link between DVs, demographics and neuropsychological performance. Cognitively healthy participants underwent a neuropsychological assessment and answered a retrospective questionnaire on NDDs symptoms in childhood and in adulthood. Using a k-means clustering based on questionnaire answers, participants were assigned to a DV+ (with DV) or a DV− (without) cluster. The final sample consisted in 84 participants [age: 69 (8); years of education: 14 (3); 57% females]. In childhood, self-management, reading and writing, school performance, and visuospatial difficulties were reported. In adulthood, difficulties were mainly in self-management. Clustering revealed a DV in 15% of individuals during childhood, and in 21% during adulthood. Throughout life, 86% of participants had a consistent cluster assignation, while 13% changed clusters between childhood and adulthood. Childhood self-management and scholar difficulties significantly predicted self-management difficulties in adulthood. Nevertheless, no link was found between DVs, demographics and neuropsychological performance. Results highlight the relevance of lifespan, dimensional investigations of NDDs; and are discussed in terms of cognitive reserve, compensation processes, heterotypic continuity and psychopathological progression.

**Public Significance Statement:** This preliminary study suggests that developmental vulnerabilities can be detected in cognitively healthy aging individuals. Difficulties in their childhood may relate to self-management, reading and writing, school performance, and visuospatial skills; and may predict self-management difficulties in adulthood.

**Highlights:**

- Lifespan neurodevelopmental vulnerabilities (DVs) were screened using a retrospective questionnaire in 84 cognitively healthy older adults.
- A data-driven dimensional analysis identified DVs in 15% of participants during childhood and 18% during adulthood.
- In childhood, reported difficulties included self-management, reading and writing, school performance, and visuospatial skills. In adulthood, difficulties were mainly in self-management.
- Childhood self-management and scholar difficulties significantly predicted self-management difficulties in adulthood.

## Introduction

The increase of life expectancy over the last century has led to a growing interest in promoting “healthy aging” and “disease-free longevity” through the identification of risk and protective factors (Garmany et al., 2021; Scott et al., 2021). While late-life proximate factors may be levers to act on, Walhovd, Lövden and Fjell underlined the importance of the lifespan influence of developmental factors on brain and cognition (Walhovd et al., 2023). Indeed, for example, more than two-thirds of the association between IQ and cortical thickness in individuals in their seventies would be explained by childhood IQ (Karama et al., 2014). However, cognitive, behavioral and affective development can be disrupted by neurodevelopmental disorders (NDDs). The growing interest in NDDs’ lifelong consequences has encouraged research into their impact during cognitive ageing (Antolini & Colizzi, 2023; Thapar et al., 2017).

NDDs are a group of syndromes with extensive repercussions on patients’ global functioning. According to the most recent classification, NDDs comprise among others, communication disorders, autism spectrum disorder (ASD), attention-deficit/hyperactivity disorder (ADHD) and specific learning disorders (SLD) (American Psychiatric Association, 2013). However, these definite categories are challenged by the frequent comorbidity between diagnoses (Bonti et al., 2024; Pennington, 2006; Thapar et al., 2017). This comorbidity is better accounted for when NDDs are viewed as a continuum of impairments in domain-specific and central dimensions of cognition, rather than as distinct entities (Demetriou et al., 2024; Dyck et al., 2011; Thapar et al., 2017). Besides, NDDs arise during childhood but their cognitive impact may extend to adulthood (e.g., American Psychiatric Association, 2013; Antolini & Colizzi, 2023; Howlin et al., 2004). For instance, it has been shown that adults with a history of language SLD had persisting language impairment and literacy difficulties in early adulthood (Whitehouse et al., 2009). NDDs also have long-lasting functional consequences, notably in adults with a history of ADHD, ASD or SLD, who may be more prone to sickness absence and work disability pension (Virtanen et al., 2020); or else in men with ADHD, who may have significantly worse educational, occupational, economic, marital and social outcomes (R. G. Klein et al., 2012) than their peers.

The use of questionnaires to detect NDDs is coherent with this dimensional, lifespan approach to NDDs (Antolini & Colizzi, 2023; Demetriou et al., 2024; Thapar et al., 2017). When used retrospectively, they can reflect atypicalities throughout the lifespan, that can further be linked with domains or dimensions of cognition. Several studies have retrospectively detected NDDs in ageing populations with the help of questionnaires (Golimstok et al., 2010; Ivanchak et al., 2011; Metzler-Baddeley et al., 2008; Rhodus et al., 2020; Seifan et al., 2018). However, if these studies made important connections between developmental factors and neurocognitive disorders (for a review, cf. Siguier et al., 2024), little is still known about neurodevelopmental vulnerabilities (DVs) in cognitive aging.

The present study thus aims at exploring DVs manifestations from childhood to adulthood in cognitive aging, with a factorial approach and a clustering analysis based on a questionnaire. This questionnaire was previously employed in a sample of individuals with neurocognitive disorders (Siguier et al., 2025). Part of the cognitive aging sample of the present study was included in this previous investigation. Our first objective was to describe domains of vulnerability linked with NDDs in a cognitively ageing population, in childhood and in adulthood. As NDDs are frequently comorbid, we expected several cognitive domains to be impaired in childhood and in adulthood. The second objective was to estimate the frequency of DVs in a cognitively ageing population. We conjectured the proportion of individuals expressing a DV would be equivalent to that previously found with this questionnaire in a smaller sample, i.e., 15% (Siguier et al., 2025). The third goal was to evaluate the persistence of symptoms of DVs from childhood to adulthood. We hypothesized an association between childhood and adulthood scores on the questionnaire. Finally, the fourth objective was to explore the link between neurodevelopmental vulnerabilities and demographic and cognitive variables.

## Method

### Standard Protocol Approvals, Registrations, and Participant Consents

Participants were prospectively recruited from February to June 2024. All provided written informed consent and the study was approved by local institutional review boards (Research Ethics Committee file No. 2023_765). The protocol was in accordance with the Helsinki Declaration of 1975, as revised in 2000.

### Transparency and Openness

As the ethical agreement did not cover the dissemination of de-identified data or codes, these are only available from the corresponding author upon reasonable request. There were no preregistrations.

### Participants

Participants had French as first language and no cognitive complaints. This was objectively measured by their MMSE score indicating no general cognitive impairment relative to their education level (Kalafat 2003). Ninety participants entered the protocol. Left-handed and ambidextrous individuals are hereinafter referred to as non-right-handed (nRH) as opposed to right-handed individuals (RH). Because this is an exploratory study, the sample size was determined based on the sample size of patients from a previous study (Siguier et al., 2025).

### Procedure

This is a cross-sectional study. Participants completed one visit of approximately 45 minutes during which their history of NDD was investigated and their current cognitive functioning was evaluated.

#### Neurodevelopmental vulnerability questionnaire

All participants answered a questionnaire evaluating life-span NDDs symptoms (cf. Sup. Figure 2 for English version) previously described (Siguier et al., 2025). The questionnaire does not provide a diagnosis, but screens for traits of NDDs described in the DSM-5 (American Psychiatric Association, 2013). It was built to reflect broad and overlapping cognitive domains such as language, mathematical thinking, coordination and attention. It was developed by the GREDEV commission (working group for the assessment of neurodevelopmental disorders in adults) as part of the GRECO (working group for cognitive assessment). The questionnaire includes 4 items of anamnesis (scored 0 if no element is reported, 1 if at least one is reported) targeting previous diagnoses of NDDs, classes repetitions, previous needs of support in primary school and previous medical or paramedical needs. These items are followed by 46 items with a Likert scale from 0 to 5 [27 for childhood (raw α=.85), e.g., “Did you find it hard to sit still? Were you told you were unruly and restless?”; 19 for adulthood (raw α=.83), e.g., “Do you have difficulties with spelling?”]. Higher scores reflect more frequent or severe difficulties. The experimenters (AO, DA, GV, PP) read all items out loud and collected participants’ answers. There was no influence of the experimenter on the total rescaled score of the questionnaire (One-way ANOVA, df=3, F=.722, p=.542).

#### Neuropsychological assessment

Participants underwent a cognitive assessment evaluating language [BECS-GRECO confrontation naming (Merck et al., 2011); ECLA16 irregular words dictation and text reading (Gola-Asmussen et al., 2011)], executive functioning [“S” lexical fluency, Go–No Go and Conflicting instructions from the FAB (Dubois et al., 2000)], visuospatial processing [Rey-Osterrieth figure copy (Osterrieth, 1944; Tremblay et al., 2015)] and anterograde memory [Rey-Osterrieth figure delayed recall (Tremblay et al., 2015)].

### Statistical analysis

Analyses were computed with R v4.4.2 (R Core Team, 2024). For all analyses, type I error threshold was set to .05.

Demographic characteristics, neuropsychological performance and scores derived from the questionnaire were compared across male and female participants. This comparison was motivated by the higher prevalence of NDDs in males than in females described in literature (American Psychiatric Association, 2013; May et al., 2019; Rutter et al., 2003).

An exploratory factor analysis (EFA) was performed on the 27 items of the childhood part of the questionnaire (package psych; Revelle, 2007). Participants were then partitioned into clusters by a k-means clustering algorithm with 100 random starting values, on the basis of the factor loadings extracted from the EFA (packages Factoextra, Kassambara & Mundt, 2020; FactoMineR, Lê et al., 2008; and cluster, Maechler et al., 2022). The number of clusters was defined according to the gap statistic and average silhouette width. The resulting clusters were characterized through comparison of factors loadings, demographical variables, neuropsychological scores and rescaled scores on each part of the questionnaire. Rescaled scores were calculated as the percentage of the maximum possible score in each part of the questionnaire. The same procedure was then independently applied on the 19 items of the adulthood part of the questionnaire.

Relationships between parts of the questionnaire and demographic variables were explored through correlations, linear modelling and Robust Linear Regression (RLM, package MASS, Venables & Ripley, 2002). RLM provides an alternative to least squares regression in the case of outliers and non-normally distributed residuals. We compared the different models obtained with backward elimination on the basis of corrected Akaike Information Criterion (AICc) and weight (package MuMin, Bartoń, 2010).

## Results

### Participants

From the 90 participants first included in the protocol, 6 were excluded from the analyses: 4 had an impaired MMSE score, 1 had Parkinson’s disease, 1 had Spanish as mother-tongue. A total of 84 subjects were thus included in the analyses. Forty-one of them were described previously (Siguier et al., 2025). The demographic characteristics of the sample are displayed in Table 1 in addition to their neuropsychological performance and the scores obtained from the questionnaire. No statistically significant difference was found between male and female participants.

**Table 1:**
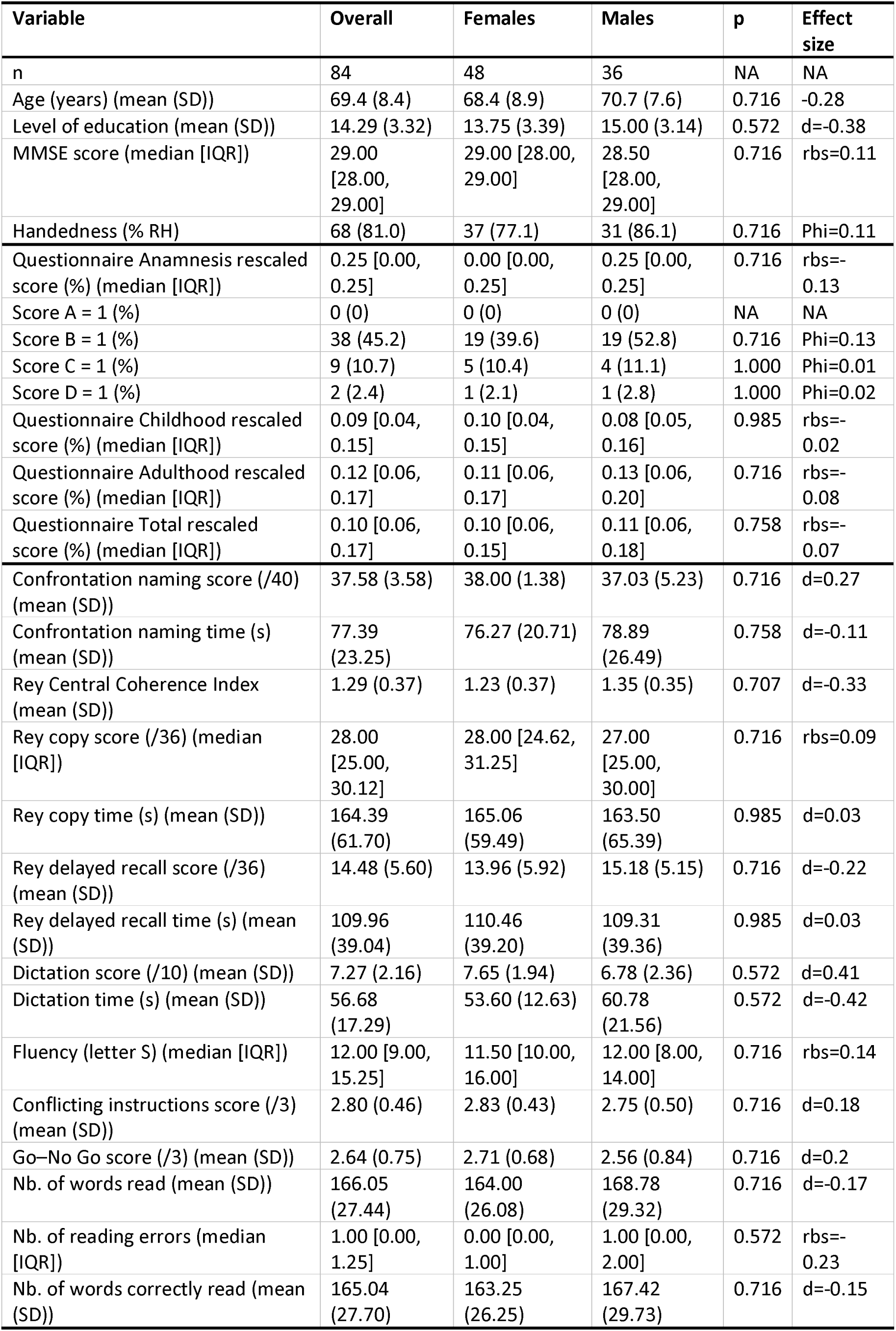
Demographics, questionnaire and cognition description of sample. P-values were calculated using only the stratified columns (males vs females) and were HDR adjusted. RH = right-handedness. Item A) “Have you been diagnosed with a neurodevelopmental disorder? If so, which one(s)?”. Item B) “Have you repeated a year and/or been in an adapted class?”. Item C) “Did you need help learning to read, write, count or draw in primary school?”. Item D) Did you receive medical or paramedical support, such as: speech and language therapy, occupational therapy, psycho-motor therapy, psychologist, child psychiatrist, neuropsychologist?”.

### Exploratory factor analysis

Principal axis extraction was used for its applicability on non-normal distributions and oblimin rotation was chosen to allow collinearity between factors. The factors were interpreted through a consensus agreement according to the cognitive or behavioral domains addressed in the contributing questions.

#### Childhood questionnaire

Bartlett’s test of sphericity indicated that the correlation structure of the EFA was adequate for factor analyses (χ²=412, df=91, p<.0001). Based on their Kaiser-Meyer-Olkin test values, 11/27 items of the childhood part of the questionnaire were kept for the EFA (general KMO = .80, cf. Sup. Table 1). The solution offering the best compromise between interpretability of factors and accounted variance (51%) was yielded by a parallel analysis with a cut-off point of .49 and 4 factors. The factors were interpreted follows: self-management (PA1), reading and writing (PA2), school repercussions (PA3) and visuospatial abilities (PA4). Results of this analysis are presented in Sup. Table 2. Correlations between factors are presented in Supplementary Table 3.

**Table 2:** Comparison of clusters based on childhood part of the questionnaire. P-values were calculated using only the stratified columns (DV– vs DV+) and were HDR adjusted. Correction of p-values was performed separately for factor loadings. RH = right-handedness. Item A) “Have you been diagnosed with a neurodevelopmental disorder? If so, which one(s)?”. Item B) “Have you repeated a year and/or been in an adapted class?”. Item C) “Did you need help learning to read, write, count or draw in primary school?”. Item D) Did you receive medical or paramedical support, such as: speech and language therapy, occupational therapy, psycho-motor therapy, psychologist, child psychiatrist, neuropsychologist?”.

| Variable | Overall | chDV- | chDV+ | p | p adjusted | Effect Size |
| --- | --- | --- | --- | --- | --- | --- |
| n | 84 | 71 | 13 |  | NA | NA |
| Age (years) (mean (SD)) | 69.37<br>(8.41) | 69.92<br>(8.23) | 66.38<br>(9.11) | 0.166 | 0.640 | d=0.23 |
| Level of education (mean (SD)) | 14.29<br>(3.32) | 14.11<br>(3.37) | 15.23<br>(3.00) | 0.267 | 0.719 | d=-0.15 |
| MMSE score (median [IQR]) | 29.00<br>[28.00,<br>29.00] | 29.00<br>[28.00,<br>29.00] | 28.00<br>[28.00,<br>29.00] | 0.532 | 0.810 | rbs=0.11 |
| Sex (% Males) | 36 (42.9) | 29 (40.8) | 7 (53.8) | 0.571 | 0.810 | Phi=0.1 |
| Handedness (% RH) | 68 (81.0) | 59 (83.1) | 9 (69.2) | 0.432 | 0.810 | Phi=0.13 |
| Questionnaire Anamnesis rescaled score (%) (median [IQR]) | 0.25<br>[0.00,<br>0.25] | 0.00<br>[0.00,<br>0.25] | 0.25<br>[0.25,<br>0.50] | 0.007 | **0.047** | rbs=-0.43 |
| Score A = 1 (%) | 0 (0) | 0 (0) | 0 (0) | NA | NA | NA |
| Score B = 1 (%) | 38 (45.2) | 28 (39.4) | 10 (76.9) | 0.028 | 0.151 | Phi=0.27 |
| Score C = 1 (%) | 9 (10.7) | 5 (7.0) | 4 (30.8) | 0.040 | 0.180 | Phi=0.28 |
| Score D = 1 (%) | 2 (2.4) | 2 (2.8) | 0 (0.0) | 1.000 | 1.000 | Phi=0.07 |
| Questionnaire Childhood rescaled score (%) (median [IQR]) | 0.09<br>[0.04,<br>0.15] | 0.07<br>[0.04,<br>0.12] | 0.25<br>[0.22,<br>0.33] | <0.001 | **0.009** | rbs=-0.95 |
| Questionnaire Adulthood rescaled score (%) (median [IQR]) | 0.12<br>[0.06,<br>0.17] | 0.09<br>[0.05,<br>0.16] | 0.26<br>[0.18,<br>0.33] | <0.001 | **0.009** | rbs=-0.8 |
| Questionnaire Total rescaled score (%) (median [IQR]) | 0.10<br>[0.06,<br>0.17] | 0.09<br>[0.05,<br>0.13] | 0.25<br>[0.23,<br>0.35] | <0.001 | **0.009** | rbs=-0.94 |
| PA1, Self-management (median [IQR]) | -0.30 [-<br>0.63,<br>0.33] | -0.48 [-<br>0.65,<br>0.07] | 1.24<br>[1.01,<br>2.53] | <0.001 | **0.001** | rbs=-0.89 |
| PA2, Reading and writing (median [IQR]) | -0.33 [-<br>0.48,<br>0.16] | -0.37 [-<br>0.50, -<br>0.14] | 0.95<br>[0.42,<br>2.12] | <0.001 | **0.001** | rbs=-0.84 |
| PA3, Scholar repercussion (median [IQR]) | -0.30 [-<br>0.46,<br>0.18] | -0.32 [-<br>0.48, -<br>0.05] | 1.57<br>[0.18,<br>1.79] | 0.001 | **0.001** | rbs=-0.61 |
| PA4, Visuo-spatial (median [IQR]) | -0.31 [-<br>0.48,<br>0.04] | -0.39 [-<br>0.50, -<br>0.15] | 0.10 [-<br>0.16,<br>0.80] | <0.001 | **0.001** | rbs=-0.63 |
| Confrontation naming score (/40) (median [IQR]) | 38.00<br>[37.00,<br>39.00] | 38.00<br>[37.00,<br>39.00] | 39.00<br>[38.00,<br>40.00] | 0.267 | 0.719 | rbs=-0.19 |
| Confrontation naming time (median [IQR]) | 73.50<br>[60.00,<br>87.00] | 73.00<br>[60.50,<br>86.50] | 74.00<br>[59.00,<br>90.00] | 0.882 | 0.953 | rbs=0.03 |
| Rey Central Coherence Index (median [IQR]) | 1.33<br>[1.07,<br>1.53] | 1.29<br>[1.09,<br>1.54] | 1.43<br>[0.81,<br>1.51] | 0.683 | 0.810 | rbs=0.07 |
| Rey copy score (/36) (mean (SD)) | 27.25<br>(4.60) | 27.40<br>(4.58) | 26.42<br>(4.85) | 0.484 | 0.810 | d=0.08 |
| Rey copy time (s) (median [IQR]) | 147.00<br>[124.50,<br>186.00] | 147.00<br>[123.00,<br>184.50] | 149.00<br>[138.00,<br>214.00] | 0.574 | 0.810 | rbs=-0.1 |
| Rey delayed recall score (/36) (mean (SD)) | 14.48<br>(5.60) | 14.49<br>(5.73) | 14.46<br>(5.06) | 0.989 | 1.000 | d=0 |
| Rey delayed recall time (s) (median [IQR]) | 101.00<br>[84.50,<br>134.50] | 100.00<br>[85.50,<br>128.00] | 115.00<br>[82.00,<br>143.00] | 0.616 | 0.810 | rbs=-0.09 |
| Dictation score (/10) (median [IQR]) | 8.00<br>[6.00,<br>9.00] | 7.00<br>[6.00,<br>9.00] | 9.00<br>[4.00,<br>9.00] | 0.515 | 0.810 | rbs=-0.11 |
| Dictation time (s) (median [IQR]) | 53.00<br>[46.00,<br>60.00] | 54.00<br>[46.50,<br>60.00] | 50.00<br>[45.00,<br>60.00] | 0.616 | 0.810 | rbs=0.09 |
| Fluency (letter S) (mean (SD)) | 12.17<br>(4.44) | 12.30<br>(4.46) | 11.46<br>(4.48) | 0.537 | 0.810 | d=0.1 |
| Conflicting instructions score (/3) (median [IQR]) | 3.00<br>[3.00,<br>3.00] | 3.00<br>[3.00,<br>3.00] | 3.00<br>[3.00,<br>3.00] | 0.293 | 0.719 | rbs=-0.12 |
| Go-No Go score (/3) (median [IQR]) | 3.00<br>[3.00,<br>3.00] | 3.00<br>[3.00,<br>3.00] | 3.00<br>[2.00,<br>3.00] | 0.206 | 0.695 | rbs=0.16 |
| Nb. of words read (mean (SD)) | 166.05<br>(27.44) | 166.56<br>(25.96) | 163.23<br>(35.62) | 0.690 | 0.810 | d=0 |
| Nb. of reading errors (median [IQR]) | 1.00<br>[0.00,<br>1.25] | 1.00<br>[0.00,<br>1.00] | 0.00<br>[0.00,<br>2.00] | 0.879 | 0.953 | rbs=-0.02 |
| Nb. of words correctly read (mean (SD)) | 165.04<br>(27.70) | 165.61<br>(26.00) | 161.92<br>(36.74) | 0.662 | 0.810 | d=0.01 |

#### Adulthood questionnaire

Bartlett’s test of sphericity indicated that the correlation structure of the EFA was adequate for factor analyses (χ²=162, df=15, p<.0001). Based on their Kaiser-Meyer-Olkin test values, 6/19 items of the adulthood part of the questionnaire were kept for the EFA (general KMO = .82, cf. Sup. Table 4). The solution offering the best compromise between interpretability of factors and accounted variance (45%) was yielded by a parallel analysis with a cut-off point of .49 and 1 factor. This factor was interpreted as reflecting ‘self-management’ difficulties (Sup. Table 5).

### Clustering

#### Childhood questionnaire

Average silhouette width indicated k=2 was the optimal number of clusters (cf. Sup. Figure 3). Cluster 1 and cluster 2 encompassed 71 (85%) and 13 (15%) individuals, respectively (Figure 1). Compared to participants in Cluster 1, participants in Cluster 2 were characterized by high scores on each part of the questionnaire and high individual loadings on every factor (cf. Table 2). These results suggested that Cluster 2 included participants with more frequent and/or severe difficulties associated with a neurodevelopmental vulnerability during childhood (chDV). Therefore, cluster 1 and cluster 2 will hereafter be named “chDV-” and “chDV+”, respectively. No difference was observed between the two clusters on demographical and cognitive variables.

**Figure 1:**
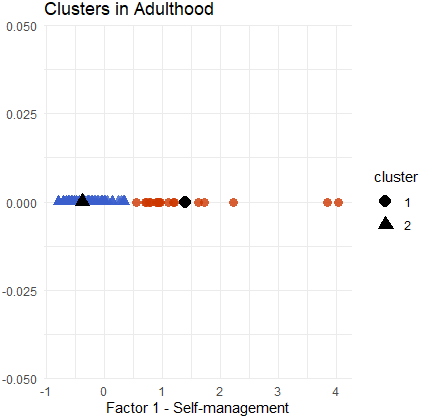
Childhood clusters visualization on the first two dimensions. Enlarged symbols indicate clusters centroids. Of note, higher scores on the questionnaire reflect more frequent or severe difficulties.

**Figure 1:**
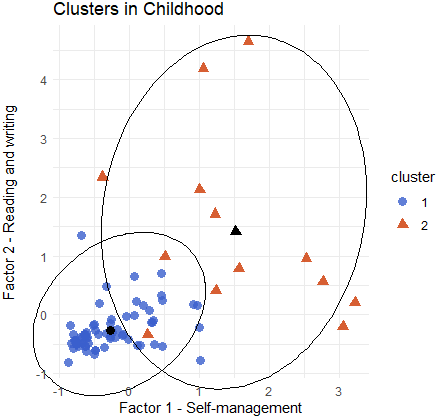
Adulthood clusters visualization on the unique dimension. Enlarged symbols indicate clusters centroids. Of note, higher scores on the questionnaire reflect more frequent or severe difficulties.

**Figure 3:**
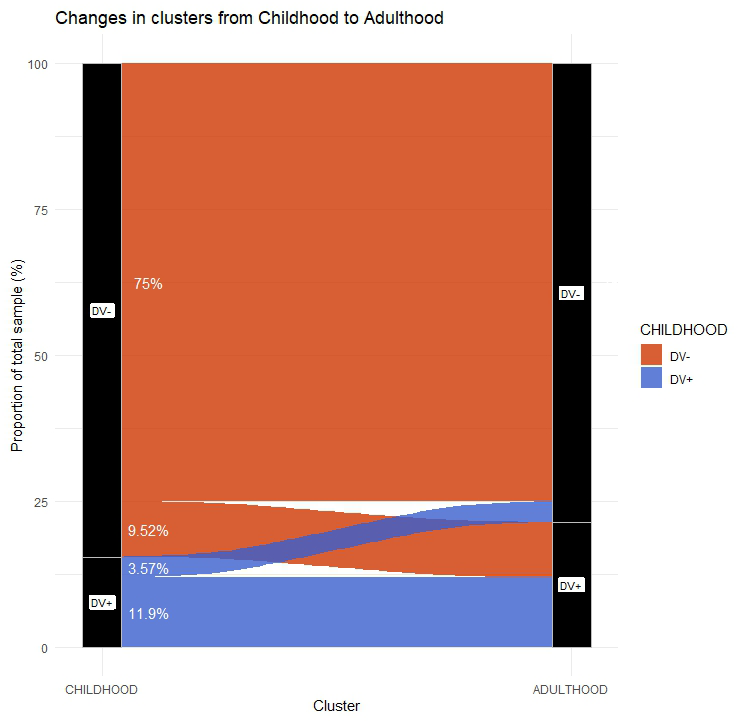
Cluster assignments in childhood and in adulthood. DV = developmental vulnerability

#### Adulthood questionnaire

Average silhouette width indicated the optimal number of clusters was k=2 (Sup. Figure 6). Cluster 1 and cluster 2 encompassed 18 (21%) and 66 (79%) individuals, respectively (Figure 2). Compared to participants in Cluster 2, participants in Cluster 1 were characterized by high scores on each part of the questionnaire, except the anamnesis. They had high individual loadings on the factor PA1 (Self-management) (Table 3). These results suggested that Cluster 1 included participants with more frequent and/or severe DV consequences during adulthood (adDV). Therefore, cluster 1 and cluster 2 will hereafter be named “adDV+” and “adDV-”, respectively. No difference was observed between the two clusters on demographical and cognitive variables.

**Table 3:**
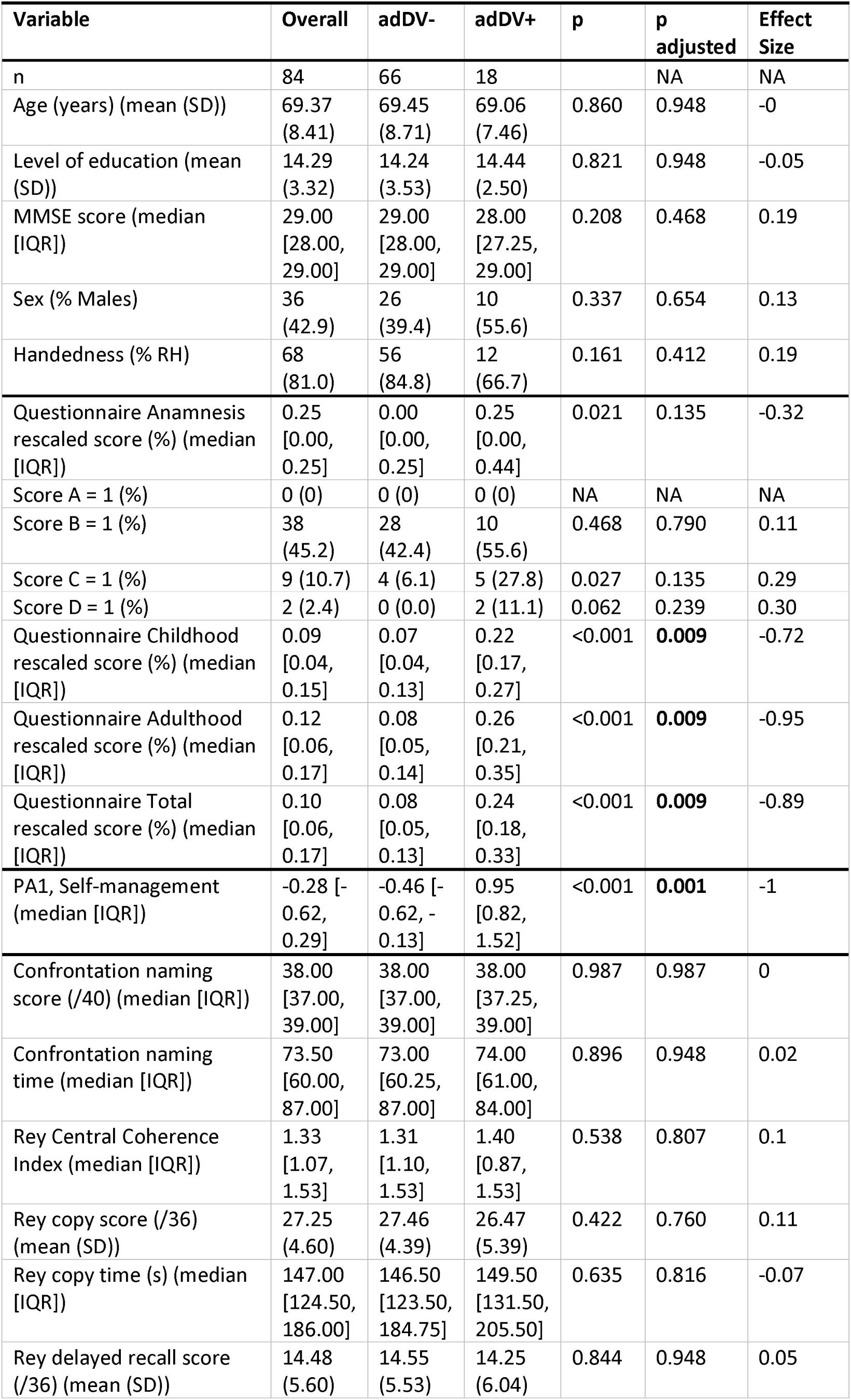
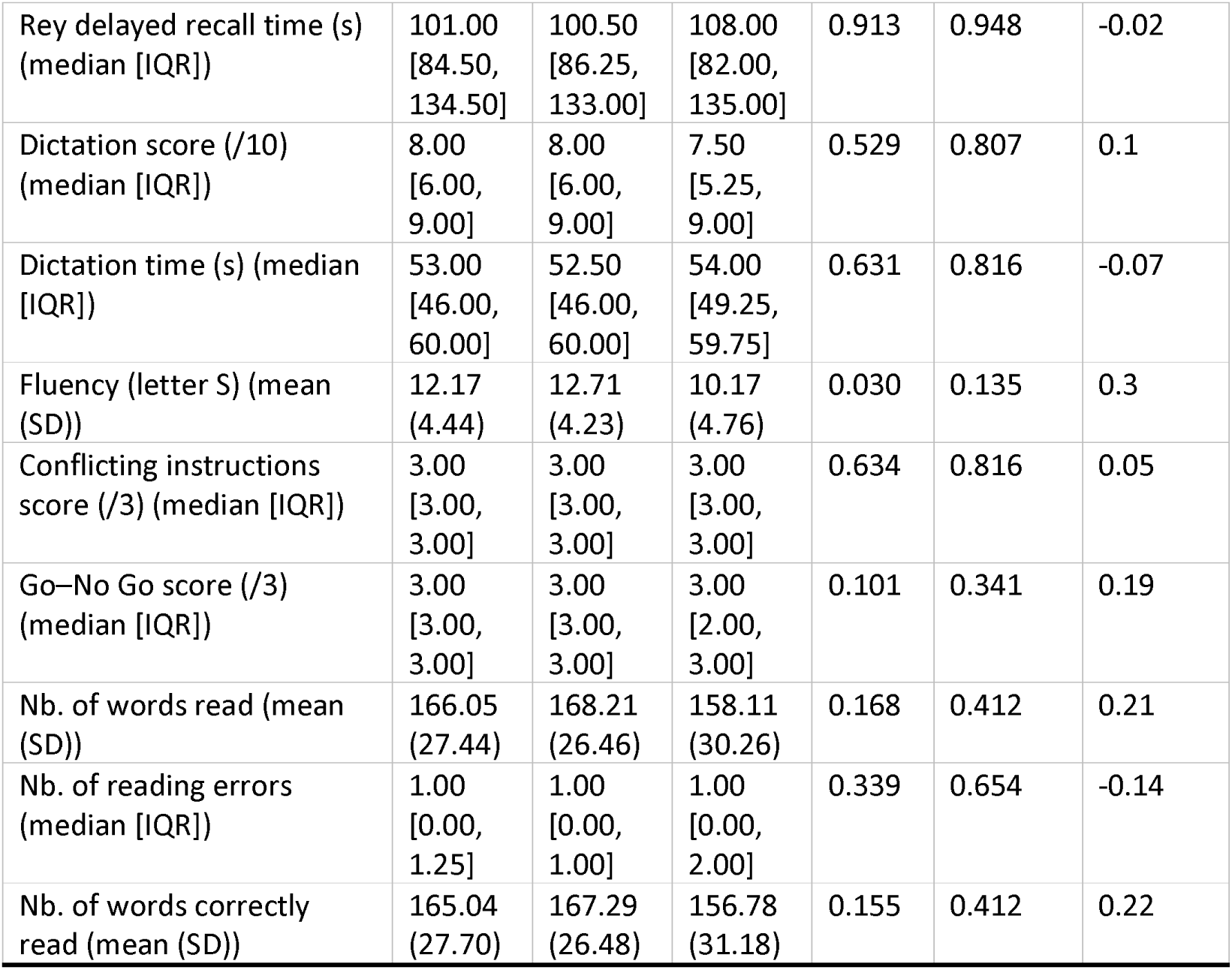
Comparison of clusters based on adulthood part of the questionnaire. P-values were calculated using only the stratified columns (DV– vs DV+) and were FDR adjusted. Correction of p-values was performed separately for factor loadings. RH = right-handedness. Item A) “Have you been diagnosed with a neurodevelopmental disorder? If so, which one(s)?”. Item B) “Have you repeated a year and/or been in an adapted class?”. Item C) “Did you need help learning to read, write, count or draw in primary school?”. Item D) Did you receive medical or paramedical support, such as: speech and language therapy, occupational therapy, psycho-motor therapy, psychologist, child psychiatrist, neuropsychologist?”.

### Lifespan trajectories

Spearman correlation indicated a positive correlation between childhood and adulthood rescaled scores (rho= .61; p<.0001).

#### Clustering from childhood to adulthood

Seventy-five percent (n=63) of the total sample were classified as DV– in both the childhood and the adulthood parts of the questionnaire (Figure 3, Sup. Table 6) while 12% (n=10) were classified as DV+ on both parts. Ten percent (n=8) were classified as DV+ only during adulthood and 4% (n=3) were classified as DV+ only during childhood.

#### Prediction of adulthood factor loading

Linear modelling and RLM were used to test if the loadings on the four childhood factors (PA 1 to 4), age, sex, education, MMS score and handedness significantly predicted adulthood factor loading extracted from the EFA.

The best model (R²= 0.50 [0.32;0.60], adjusted R²=0.48, RSE = 0.66 on 80 df) was a linear model: Adulthood factor loading = –6.82 x 10 + 0.58*(PA1) – 0.14*(PA2) + 0.56*(PA3). Loadings on the first and third childhood factors (PA1-Self-management and PA3-Scholar repercussion) were the only significant predictors throughout all tested models (p<0.0001 in this model).

## Discussion

This preliminary, data-driven study explored lifespan dimensions of neurodevelopmental vulnerabilities (DV) in cognitive aging. This was done via a retrospective questionnaire screening for DVs from childhood to adulthood. This questionnaire was previously employed in the context of Alzheimer’s disease and frontotemporal dementia, as well as in a fraction of the sample of healthy individuals presented here (Siguier et al., 2025). This work builds on a field of literature relying on questionnaires to retrospectively screen for NDDs. Several studies have indeed done so in ageing populations (Golimstok et al., 2010; Ivanchak et al., 2011; Metzler-Baddeley et al., 2008; Rhodus et al., 2020; Seifan et al., 2018). Nevertheless, when they were validated, the main limit of the questionnaires employed remained their focus on specific categories of NDDs, such as ADHD (Ivanchak et al., 2011) or neurodevelopmental learning and attentional disorders (Seifan et al., 2018). Instead, we relied on a questionnaire for its ability to capture a relatively wide range of symptoms, and thus to transcend labels of NDDs (Colvin & Sherman, 2020; Magnin et al., 2023). This allowed a more comprehensive approach of NDDs, respective of their potential overlap.

In the overall sample, the corrected score in the childhood and adulthood parts of the questionnaire were only of 9% and 12%, respectively. This reveals that in this sample, DVs-related difficulties were probably not particularly frequent, pervasive or intense. This is consistent with the fact that no participant had received a formal diagnosis of NDD. This may be due to the selection of individuals without cognitive complaint, with a fairly high level of education. However, this might equally reflect the dearth of screening of NDDs in generations born before the 1980’s, as the term “Developmental disorders” was first coined in the DSM-III (American Psychiatric Association, 1980; Morris-Rosendahl & Crocq, 2020). Therefore, the low frequency (2.4%) of reported medical or paramedical support during childhood in our sample appears unsurprising. The repetition of at least a class was, on the contrary, more common (45.2% of positive answers). This would appear coherent considering that among the four childhood factors that emerged from the questionnaire, one reflected scholar repercussions and another reflected reading and writing. Nonetheless, only 10.7% of participants reported they had needed help learning to read, write, count or draw in primary school. This discrepancy shows that academic struggles (e.g., class repetitions) cannot be the sole proxy for NDDs in an aging population. This is further emphasized by the similar level of education between DV+ and DV– clusters.

The first objective of this study was to describe NDDs-related domains of vulnerability by means of an exploratory factorial analysis. It is important to underline that the resulting factors’ labels are subject to interpretation and cannot exhaustively match all items loading to a given factor. In childhood, four domains emerged: self-management, reading and writing, scholar repercussions, and visuospatial abilities. These would reflect core domains of NDDs, close to the dimensions described by (Demetriou et al., 2024; Thapar et al., 2017). Their collinearity, low but existent, is in line with the overlap of multiple NDDs dimensions we hypothesized. In adulthood, contrary to our assumption, only one domain emerged. It seemed to reflect daily emotional and cognitive self-management difficulties also encompassed in the childhood’s eponym factor, but transposed into adulthood-specific situations. Difficulties in other domains may indeed be less likely to be reported in adulthood, either because of a true remission, either because impairments in these domains have been efficiently compensated. The other possibility is that these difficulties would be captured if individuals would not self-select their environment to match their strengths. This phenomenon is referred to as “niche construct” or “environmental accommodation” (Johnson et al., 2015; Thapar et al., 2017).

The second objective of this study was to estimate the frequency of DVs in a cognitively aging population. The reader should keep in mind that this study is not epidemiologic. The findings are preliminary and should thus be interpreted with caution due to limited power. In our sample, the frequency of DVs in childhood was estimated to 15%. This was expected since in a subset of the present sample, this frequency was 14.6% (Siguier et al., 2025). Both estimations appear to exceed worldwide prevalence rates (Antolini & Colizzi, 2023; Olusanya et al., 2023). However, prevalence rates in childhood are classically given for specific categories of NDDs (e.g., 5% for ADHD; 5% for SLDs; 1.5% for ASD) and vary considerably across methodological and social contexts (Antolini & Colizzi, 2023; Francés et al., 2022; Olusanya et al., 2023). Here, the focus was on the dimensions related to NDDs, and the categories can therefore be considered to be confounded. Besides, the questionnaire screens for traits of NDDs, which are by essence more frequent than definite diagnoses (Thapar et al., 2017). The same conclusion can be drawn for the estimated 21% of adulthood DVs in our sample. Here again, prevalence rates among adults are rarely estimated independently of categories (e.g., 4% for ADHD; 1–2.5% for SLDs; 1% for ASD) (Virtanen et al., 2020). Besides, estimates in adulthood vary according to which diagnosis criteria are applied, as NDDs features can change throughout development. In ADHD for example, the persistence rate varies from ∼15% at age 25 years when full criteria for ADHD are considered, to ∼65% when cases in partial remission are included (Faraone et al., 2006, 2021). Importantly, the DV proportion found in adulthood in our sample cannot only reflect long-lasting difficulties, as it is superior to that found in childhood. This could be explained by a possible recency effect: difficulties in adulthood may be vividly perceived and thus receive higher scoring; as opposed to earlier struggles from childhood that may be minimized. Likewise, difficulties could be judged significant only in adulthood, when increasing demand exceeds limited capacities (Livingston & Happé, 2017). Of course, the adult onset of NDDs, as suggested for certain cases of ADHD notably (Caye et al., 2017; Moffitt et al., 2015), cannot be excluded. This hypothesis is supported by the significant difference in the anamnesis scores found between the DV– and DV+ groups in childhood, but not in adulthood. This observation suggests that at least certain individuals of the adDV+ group did indeed not require earlier support in childhood. However, why and how the early manifestations of these cases would be suppressed is still an open question (Thapar et al., 2017). Finally, it is possible that the adulthood self-management factor does not specifically reflect NDDs-related traits. It could also represent different disorders, such as psychiatric disorders. It has indeed been proposed that NDDs and psychiatric disorders would constitute a continuum, and not separate entities (Antolini & Colizzi, 2023).

The third objective was to evaluate the persistence of symptoms of DVs throughout the lifespan. Here again, it is worth noting that interpretations are theoretical because of the small subsamples’ sizes. Eighty-seven percent of participants belonged to the same cluster in childhood and in adulthood. Among them, the majority (n=63, 75%) were classified as DV– in both parts of the questionnaire. The others (n=10, 12%) were classified as DV+, suggesting persisting DV repercussions from childhood to adulthood. Indeed, as mentioned earlier, NDDs symptoms may decline but repercussions and diagnoses may persist during adulthood (e.g., (American Psychiatric Association, 2013; Antolini & Colizzi, 2023; Howlin et al., 2004; R. G. Klein et al., 2012; Virtanen et al., 2020; Wang et al., 2024; Whitehouse et al., 2009). Fourteen percent of participants, on the contrary, had discordant clusters’ assignations throughout life. As the clustering directly depended on factors, interpretation of these changes is closely related to assumptions previously formulated about factors found in childhood and in adulthood. Therefore, the three individuals (4%) classified as DV+ only during childhood could exemplify either processes of change in core symptoms, environmental accommodation, compensation or remission, as detailed earlier (Demetriou et al., 2024; Johnson et al., 2015; Livingston & Happé, 2017; Thapar et al., 2017). Conversely, the eight participants (10%) classified as DV+ only during adulthood (Caye et al., 2017; Thapar et al., 2017) could be concerned with late-onset NDDs, or with independent traits resulting from a lack of specificity of the adulthood items.

We also investigated the link between symptoms of DVs throughout the lifespan with a continuous approach, without dichotomizing the concept of DV. We showed that more frequent or important difficulties in self-management and in school during childhood predicted more frequent or important difficulties in self-management in adulthood. This corroborates findings from Copeland and colleagues (2015) demonstrating that individuals with a childhood psychiatric disorder were more likely to experience adverse outcomes related to health, the legal system, personal finances, and social functioning in adulthood. This was also true for participants with subthreshold psychiatric problems. They showed the best diagnostic predictor of adverse outcomes in adulthood was indeed the cumulative burden of psychopathology in childhood.

The fourth goal was to explore the link between DVs, and demographic and cognitive variables. First, sex was not associated with different scores on the questionnaire. This replicates the observation made in Siguier et al., 2025, that differs from a commonly acknowledged higher prevalence of NDDs in males than in females (American Psychiatric Association, 2013; Antolini & Colizzi, 2023; Faraone et al., 2021; Lai et al., 2015). As previously postulated (Lai et al., 2015), this could suggest that if they were systematically screened, women might be found to be as affected by NDDs as men. Second, in our continuous approach, demographic variables were not significant predictors of adulthood factor loadings. While a veritable absence of effect is possible, it may also be due to the lack of variability in these predictors in our relatively uniform sample. This also applies to the absence of significant difference in demographic variables and neuropsychological scores between the DV+ and DV-groups throughout life (an observation also made in our previous work; Siguier et al., 2025). Interpretation is limited by a potential lack of power and of exhaustiveness of the neuropsychological assessment. Indeed, our sample is free from cognitive complaints: the difference in cognitive scores between DV+ and DV-groups, if it exists, might have been too subtle to objectivize. Besides, factors extracted from the questionnaire describe general cognitive domains, as opposed to neuropsychological scores that measure more specific cognitive processes. A theory-driven data reduction applied to the questionnaire might have revealed more straightforward relationships between neuropsychological scores and DV statuses. Still, it is conceivable that cognitive impairments resulting from a childhood DV have been compensated and were thus not observable in late adulthood. More generally, self-perceived difficulties are not always quantifiable through standardized tests (C. B. Klein et al., 2023). Eventually, this absence of significant difference indicates that the clustering was not driven by demographic or cognitive characteristics, and thus that these were unlikely to bias answers to the questionnaire.

This preliminary study has several limitations. First, the reduced sample size led several items from the questionnaire to be dropped from the factorial analysis because of low sample adequacy. It is thus possible that in a different population, emerging factors would reflect slightly different dimensions. The sample size and associated reduced power may also have prevented detecting small differences between groups, especially in terms of demographic and cognitive variables. As previously discussed though, it is also possible that the absence of difference was due to a selection bias, as our overall sample was highly educated and without cognitive complaint. Indeed, in order to rule out neurodegenerative diseases, individuals with cognitive complaints were not included, which potentially also excluded individuals with DVs. Second, the comparison of DV– and DV+ groups we performed implied a dichotomization of the concept of developmental vulnerabilities. This raises the question: “Where does the cutoff point on a dimension lie?” (Thapar et al., 2017, p. 4) and, more generally, of where does the concept of “normality” ends (Canguilhem, 2013). As underlined by Thapar et al., 2017), getting a diagnosis is associated both with benefits and psycho-social risks that must not be neglected. Even if the purpose of the questionnaire was not to provide a diagnosis, we addressed these questions by defining the threshold between the DV+ and the DV-over multiple dimensions, without a priori. Besides, we also proposed a complementary, continuous approach to DVs that might reflect more accurately the clinical reality. Third, the questionnaire we employed requires a formal validation. Especially, it appears that external validity needs to be improved, particularly in regard to cognitive performance. Yet, it is possible that a more exhaustive neuropsychological assessment is required to capture this link. Besides, the adulthood part of the questionnaire could be enriched with more diverse items, in an attempt to describe more thoroughly other potential dimensions, such as visuo-spatial difficulties. Nevertheless, this study already provides encouraging indicators about the questionnaire’s validity. Face validity is at least partly insured by the fact that it was designed by experts of NDDs lifespan trajectories (Mokkink et al., 2010). Besides, Cronbach’s alphas for the childhood and adulthood parts of the questionnaire indicated good consistencies (childhood: α=0.85; adulthood: α=0.83). Regarding the factors extracted, consistency could be improved but already showed reliable overall results. In this sense, a confirmatory factorial analysis will be required to insure the construct validity of the questionnaire. The forthcoming validation should be carried in a multicentric cohort including individuals having received a formal diagnosis of NDD; and being more diverse in terms of, notably, levels of education and cognitive functioning. Diverse ethnicities should also be included, as NDD characterization is still lacking across culturally diverse samples (Olusanya et al., 2023) although diagnoses of NDDs are based on clinical features that are influenced by cultural values (for a review, see Norbury & Sparks, 2013).

All in all, this study requires replication before robust, generalizable conclusions can be drawn about the prevalence of NDDs, the dimensions and the persistence of lifespan difficulties in cognitive aging. Nevertheless, it encourages future works investigating NDDs in a dimensional manner. It represents a first step to develop a retrospective screening tool for NDDs in adults, beyond categories, and considering that NDDs are more than their defining symptoms (Thapar et al., 2017). It also emphasizes the importance of adopting a lifespan approach to NDDs, as this is likely to inform on prognosis, but also on the ‘heterotypic continuity’ and the ‘psychopathological progression’ concepts (Rutter et al., 2006). The former postulates no change in the disorder as such, but behavioral expressions varying with age (e.g., a written language disorder may manifest itself early on through difficulties with reading, and later through difficulties with spelling). The latter questions the existence of a common disorder underlying behavioral features with less “self-evident” link (e.g., progression from ADHD to a later antisocial personality disorder) (Rutter et al., 2006, p. 288). Deciphering these processes will likely require longitudinal studies integrating measurements across multiple levels (i.e., genes, molecules, cells, nerve circuits, physiology, behavior and self-reports). This is, for example, what the Research Domain Criteria (RDoC) framework proposes (Vilar et al., 2019). In this context, more research on cognitive aging is needed, as it might also shed light on the processes underlying the impact of NDDs on late-life conditions such as neurocognitive disorders (e.g., (Levine et al., 2023; Miller et al., 2018; Rutter et al., 2006; Vivanti et al., 2021; Wang et al., 2024; Yin et al., 2025).

## Data Availability Statement

The data that support the findings of this study are available from the corresponding author upon reasonable request.

## Ethics approval statement

All participants provided written informed consent and the study was approved by local institutional review boards (Research Ethics Committee file No. 2023_765). The protocol was in accordance with the Helsinki Declaration of 1975, as revised in 2000.

## Acknowledgements

We thank all members of the GREDEVad (Groupe de réflexion sur l’évaluation des troubles neurodéveloppementaux de l’adulte) of the GRECO for their contribution. Our warmest thanks go to participants and their families.

