## Supplementary material for "Neurodevelopmental Vulnerability in Cognitive Aging: A Dimensional Approach": Supp.Material

Appendices

Neurodevelopmental disorders screening questionnaire

**NAME:**

**First name:**

**Date of Birth:**

**Sex:**

- Female
- Male

**Have you been diagnosed with a neurodevelopmental disorder? If so, which one(s)?**

**Education**

1) Have you repeated a year and/or been in an adapted class?

- Yes
- No

If yes, please specify:

2) Did you need help learning to read write, count or draw in primary school?

- Private tutoring with a teacher
- Frequent practice with parents or close adults
- Special needs assistant, Individual Educational Success Plan, Tailored program, etc.?
- Part-time
- Other adaptations

3) Did you receive medical or paramedical support, such as:

- Speech and language therapy
- occupational therapy
- psycho-motor therapy
- psychologist
- child psychiatrist
- neuropsychologist

4) What is your highest level of education or most recent diploma?

5) What do you do for a living?

6) Are you:

- right-handed
- left-handed
- ambidextrous

***For each of the following questions, rate the intensity of the behavior mentioned between 0 (no difficulty) and 5 (extremely marked or frequent difficulty):***

***Part 1: When you were a child:***

1) Did you have school phobia? Did you hate school? Was it extremely difficult or even impossible to attend?

0	1	2	3	4	5

2) Did you reverse letters (e.g., writing 'hosre' instead of 'horse') or numbers (e.g., '58' instead of '85')?

0	1	2	3	4	5

3) Were you awkward or uncomfortable moving in space?

0	1	2	3	4	5

4) Did you have trouble managing your emotions (e.g., anger outbursts) or stress?

0	1	2	3	4	5

5) Did you have issues with eating, such as selective eating or an eating disorder?

0	1	2	3	4	5

6) Did you struggle finding your bearings or keeping track of time?

0	1	2	3	4	5

7) Did you have trouble remembering or learning new things?

0	1	2	3	4	5

8) Did you have difficulties in reading, being slower or making more mistakes than your peers?

0	1	2	3	4	5

9) Did you feel you were different from the others, out of step?

0	1	2	3	4	5

10) Did those around you tell you that you were inattentive, daydreaming, or that you lacked concentration?

0	1	2	3	4	5

11) Have you encountered any difficulties in your relationships with family, friends or school mates (harassment, etc.)?

0	1	2	3	4	5

12) Did you want to be the first to answer questions, not waiting for the end of the questions or interrupting?

0	1	2	3	4	5

13) Did you have difficulties with spelling (dictation)?

0	1	2	3	4	5

14) Did you find it difficult to organize your daily routine (homework, tidying up, etc.)?

0	1	2	3	4	5

15) Did you have trouble taking notes in class?

0	1	2	3	4	5

16) In primary and secondary school, did you have any difficulties with mental arithmetic (i.e., without using your fingers to count)?

0	1	2	3	4	5

17) Did you have difficulties acquiring speech or fluency: learning to speak, stuttering, articulation or pronunciation difficulties?

0	1	2	3	4	5

18) Did you struggle mastering certain gestures like buttoning clothes, tying shoelaces, brushing your teeth, using scissors, holding cutlery?

0	1	2	3	4	5

19) Did you have trouble learning your multiplication tables?

0	1	2	3	4	5

20) Did you have trouble learning to write and form letters?

0	1	2	3	4	5

21) Did you have trouble finding your bearings in space?

0	1	2	3	4	5

22) Did you have comments about the “poor appearance” of your notebooks, homework or assignments?

0	1	2	3	4	5

23) Did you find it hard to sit still? Were you told you were “unruly and restless”?

0	1	2	3	4	5

24) In primary and secondary school, did you have any difficulties with geometry (drawing figures, etc.)?

0	1	2	3	4	5

25) Have you had trouble learning to swim, play with a ball, play with a racket, rollerblade or ride a bike?

0	1	2	3	4	5

26) Did you make any careless mistakes?

0	1	2	3	4	5

27) Did you find it difficult to make friends, fit in with your classmates, or take part in extracurricular activities?

0	1	2	3	4	5

***Part 2: As an adult (before the first symptoms of neurodegenerative disease):***

***For each of the following questions, rate the intensity of the behavior mentioned between 0 (no difficulty) and 5 (extremely marked or frequent difficulty):***

28) Are you experiencing difficulties in your professional relationships?

0	1	2	3	4	5

29) Do you tend not to tie/untie your shoe laces?

0	1	2	3	4	5

30) Are you less comfortable than your peers when it comes to reading? For example, do you find it difficult to read movie subtitles?

0	1	2	3	4	5

31) Do you have trouble completing two tasks at the same time?

0	1	2	3	4	5

32) Do you consider yourself “unstable” in romantic relationships?

0	1	2	3	4	5

33) Do you have trouble understanding complex verbal instructions?

0	1	2	3	4	5

34) Do you have trouble putting things in order when doing something that requires organization?

0	1	2	3	4	5

35) Do you have trouble managing your emotions?

0	1	2	3	4	5

36) Do you consider yourself “unstable” professionally (for example, do you frequently change jobs)?

0	1	2	3	4	5

37) Do you find it difficult to finalize the last details of a project once you have achieved the most rewarding parts?

0	1	2	3	4	5

38) Do you have difficulties understanding others' emotions, humor, or implied meanings?

0	1	2	3	4	5

39) Do you have difficulties with spelling?

0	1	2	3	4	5

40) Do you have trouble remembering appointments or obligations?

0	1	2	3	4	5

41) Do you have trouble taking notes, in meetings for example?

0	1	2	3	4	5

42) Do you struggle to eat neatly if you're not paying attention?

0	1	2	3	4	5

43) Do you ever feel overly active and compelled to do something, as if you were driven against your will by a motor?

0	1	2	3	4	5

44) When something requires a lot of thought, do you ever avoid doing it or put it off?

0	1	2	3	4	5

45) Do you ever fidget or wiggle your hands or feet when you have to sit still for long periods?

0	1	2	3	4	5

46) Do you misplace items (keys, documents, mobile phone, etc.?)

0	1	2	3	4	5

**CHILD SCORE:**

**ADULT SCORE:**

**TOTAL SCORE:**

Supplementary Figure 2: English version of the questionnaire investigating retrospective neurodevelopmental disorders symptoms.

| **Items** | **MSA** |
| --- | --- |
| **General** | **.80** |
| score_1 | .81 |
| score_4 | .82 |
| score_7 | .83 |
| score_8 | .88 |
| score_10 | .81 |
| score_11 | .74 |
| score_12 | .52 |
| score_14 | .75 |
| score_15 | .64 |
| score_20 | .74 |
| score_21 | .55 |
| score_22 | .68 |
| score_24 | .59 |
| score_26 | .81 |

Supplementary Table 1: Measure of Sampling Adequacy (MSA) for the childhood part of the questionnaire

Kaiser-Meyer-Olkin test for Sampling Adequacy. Scores refer to items of the childhood part of the questionnaire.

| **Variable** | **Uniqueness** | **Loading** | **Factor** | **Raw Cronbach alpha [Feldt 95IC]** | **Cumulative Variance** | **Factor interpretation** |
| --- | --- | --- | --- | --- | --- | --- |
| **score_4**  Did you have trouble managing your emotions (e.g., anger outbursts) or stress? | 0.499 | 0.624 | PA1 | .74 [.64  .82] | 0.163 | Self-management |
| **score_10**  Did those around you tell you that you were inattentive, daydreaming, or that you lacked concentration? | 0.313 | 0.590 |  |  |  |  |
| **score_11**  Have you encountered any difficulties in your relationships with family, friends or school mates (harassment, etc.)? | 0.677 | 0.597 |  |  |  |  |
| **score_14**  Did you find it difficult to organize your daily routine (homework, tidying up, etc.)? | 0.561 | 0.690 |  |  |  |  |
| **score_8**  Did you have difficulties in reading, being slower or making more mistakes than your peers? | 0.615 | 0.555 | PA2 | .73 [.62  .82] | 0.309 | Reading and writing |
| **score_20**  Did you have trouble learning to write and form letters? | 0.281 | 0.859 |  |  |  |  |
| **score_22**  Did you have comments about the “poor appearance” of your notebooks, homework or assignments? | 0.432 | 0.633 |  |  |  |  |
| **score_7**  Did you have trouble remembering or learning new things? | 0.422 | 0.567 | PA3 | .62 [.42  .76] | 0.413 | School repercussion |
| **score_15**  Did you have trouble taking notes in class? | 0.452 | 0.717 |  |  |  |  |
| **score_21**  Did you have trouble finding your bearings in space? | 0.287 | 0.835 | PA4 | .69 [.51  .80] | 0.512 | Visuo-spatial abilities |
| **score_24**  In primary and secondary school, did you have any difficulties with geometry (drawing figures, etc.)? | 0.554 | 0.654 |  |  |  |  |

Supplementary Table 2: Exploratory Factor Analysis of the childhood items of the questionnaire.

Extraction method: Principal axis; Rotation method: Oblimin. Items designate the number of the question found in the questionnaire. Items 1, 2, 3, 5, 6, 9, 12, 13, 16, 17, 18, 19, 23, 25, 26, 27 were not included in the interpretation of factors (loadings < .49).

| **Factors** | **PA1** | **PA2** | **PA3** | **PA4** |
| --- | --- | --- | --- | --- |
| PA1 | 1.000 | 0.369 | 0.243 | 0.310 |
| PA2 | 0.369 | 1.000 | 0.171 | 0.077 |
| PA3 | 0.243 | 0.171 | 1.000 | 0.310 |
| PA4 | 0.310 | 0.077 | 0.310 | 1.000 |

Supplementary Table 3: Inter-factor correlations on the childhood part of the questionnaire.

| **Items** | **MSA** |
| --- | --- |
| **General** | **.82** |
| score_31 | 0.82 |
| score_33 | 0.81 |
| score_34 | 0.84 |
| score_35 | 0.83 |
| score_37 | 0.81 |
| score_44 | 0.80 |

Supplementary Table 4: Measure of Sampling Adequacy (MSA) for the adulthood part of the questionnaire

Kaiser-Meyer-Olkin test for Sampling Adequacy. Scores refer to items of the adulthood part of the questionnaire.

| **Variable** | **Uniqueness** | **Loading** | **Factor** | **Raw Cronbach alpha [Feldt 95IC]** | **Proportion of Variance explained** | **Factor interpretation** |
| --- | --- | --- | --- | --- | --- | --- |
| **score_31**  Do you have trouble completing two tasks at the same time? | 0.502 | 0.706 | PA1 | .82 [.75  .87] | 0.45 | Self-management |
| **score_33**  Do you have trouble understanding complex verbal instructions? | 0.476 | 0.724 |  |  |  |  |
| **score_34**  Do you have trouble putting things in order when doing something that requires organization? | 0.508 | 0.701 |  |  |  |  |
| **score_35**  Do you have trouble managing your emotions? | 0.700 | 0.548 |  |  |  |  |
| **score_37**  Do you find it difficult to finalize the last details of a project once you have achieved the most rewarding parts? | 0.550 | 0.671 |  |  |  |  |
| **score_44**  When something requires a lot of thought, do you ever avoid doing it or put it off? | 0.567 | 0.658 |  |  |  |  |

Supplementary Table 5: Exploratory Factor Analysis of the adulthood items of the questionnaire.

Extraction method: Principal axis; Rotation method: Oblimin. Items designate the number of the question found in the questionnaire. Items 28, 29, 30, 32, 36, 38, 39, 40, 41, 42, 43, 45, 46, were not included in the interpretation of factors (loadings < .49).

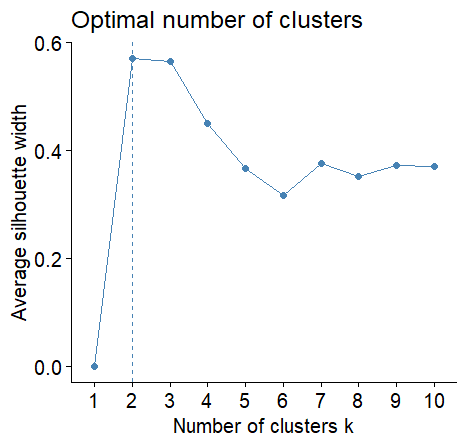

Supplementary Figure 3: Index plot showing the optimal number of clusters on the childhood part of the questionnaire.

Average silhouette width indicated k=2 was the optimal number of clusters.

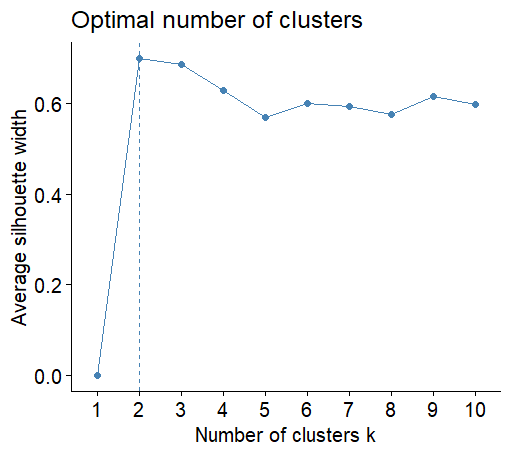

Supplementary Figure 6: Index plot showing the optimal number of clusters on the adulthood part of the questionnaire.

Average silhouette width indicated k=2 was the optimal number of clusters.

| **Childhood** | **Adulthood** | **n** | **Proportion of total sample (%)** |
| --- | --- | --- | --- |
| chDV- | adDV- | 63 | 75.00 |
| chDV+ | adDV+ | 10 | 11.90 |
| chDV- | adDV+ | 8 | 9.52 |
| chDV+ | adDV- | 3 | 3.57 |

Supplementary Table 6: Cluster assignment in childhood and in adulthood.
